# The association of UV with rates of COVID-19 transmission and deaths in Mexico: the possible mediating role of vitamin D

**DOI:** 10.1101/2020.05.25.20112805

**Authors:** M Skutsch, C Dobler, M.B.B McCall, A Ghilardi, M Salinas, M.K McCall, G Fenner Sanchez

## Abstract

The first COVID-19 case in Mexico was confirmed on 26 February 2020 and by May 3 the number of registered cases had risen to 30,927. However the rate of transmission varied greatly from city to city. We used data on temperature, humidity and ultraviolet radiation (UV) from 45 cities all over the country to explore whether there was an association between these variables and rates of transmission and rates of accumulation of COVID-19 ascribed deaths. The advantage of an in-country study of this kind is that many of the variables that can confound international studies are held constant (e.g. public health policies, methods of reporting, cultural, behavioural and genetic factors). Although the official statistics undoubtedly greatly underestimate the situation in Mexico due to lack of testing, they are underestimated in all cities so this should not introduce bias across the sample. We found that temperature and humidity had no discernible association with transmission rates but that UV during the transmission period was negatively correlated with rates of transmission, suggesting a sterilizing effect. UV in the January preceding the epidemic had a slightly higher association with transmission rates than UV during the transmission period itself. We also found negative associations of UV in the transmission period and in January with rate of cumulative deaths, but at lower levels of statistical significance. We conclude that in addition to a sterilizing effect during the transmission period, UV may have a physiological effect in reducing transmission and deaths due to COVID-19, most likely through the medium of vitamin D production in the body. This follows the growing body of medical evidence that vitamin D deficiency is associated with severity of COVID-19. However, we also found a negative correlation between altitude and rates of COVID-19 transmission, distinct and independent of the UV effect, which may indicate that other physiological processes are also present. In a multiple regression, altitude and UV together accounted for 18% of the variation in transmission rates between cities.

## Background

The future course of the on-going coronavirus (COVID-19) pandemic remains uncertain, particularly whether this may be (temporarily) mitigated in the northern hemisphere with the advent of summer. It is well established that viral respiratory infections such as influenza (Lofgren et al. 2007) and respiratory syncytial virus (Mizuta et al. 2013) follow a seasonal pattern (Cannell et al. 2008; Price et al. 2019), with peak transmissions during winter months, which could be due to climatic and meteorological factors or to social behaviour such as indoor confinement and crowding. Aldridge et al. (2020) found that the infection rate for a number of other coronaviruses in England was much higher in the winter months although there was some transmission in the summer. The World Health Organization nevertheless still cautions against assuming that summer will reduce the impact of the new corona virus (WHO, 2020).

Jüni et al. (2020) in an inter-country study found no evidence that temperature or latitude had an impact on COVID infection rates, although they did find a weak relationship with humidity; instead they found that public health measures explain much of the variation. Davies et al. (2020) compile evidence that latitude is a major controlling factor on COVID-19. Heneghan and Jefferson (2020) comparing countries found latitude had an association with COVID-19 but temperature did not. Wan et al. (2020) using global datasets have suggested that low temperatures were associated with COVID-19 spread in the early period and that in later phases higher temperature reduced this. Merow and Urban (2020) compared infection rates of COVID-19 across a number of countries and found that 36% of the variation in early incidence could be explained by the suite of factors considered, of which about half (17%) was attributable to weather or demographic factors. UV radiation was the most influential variable among these.

The difficulty of making cross country comparisons with a view to understanding the impacts of factors such as temperature, humidity and UV is that so many other conditions vary across the sample. This includes the date at which social distancing measures and testing were put in place after the first infections and also how the incidence of infection, disease and death is recorded and reported. Specific socio-economic/cultural/behavioural and genetic factors probably also play a large part (Evans, 2013), as may the age structure of the population (Lin, Yiu and Kammen 2020). By focusing on the differential rates of spread of COVID-19 in different cities within one country, Mexico, many of these other confounding factors may be minimized.

The first COVID-19 case in Mexico was detected on February 26 2020 in Mexico City and by 3 May there had been 30,927 cases confirmed and 3,332 deaths according to official data sources. The indications are however that the rate of growth of COVID-19 cases in Mexico, at least over these first ten weeks, has been slower than that observed in Italy, Spain and the USA (Figure 1). An obvious explanation for the apparently slow growth in number of cases in Mexico is that there is lack of testing and hence under-reporting. Under-reporting occurs to varying degrees in every country in the world as many cases are mild or asymptomatic and testing was not always started early, but Mexico has especially low levels of testing. On May 8, Mexico was testing 0.04 persons per 1000, while the average in OECD countries by April 28 was 22/1000 (Our World in Data, 2020).

**Figure 1:**
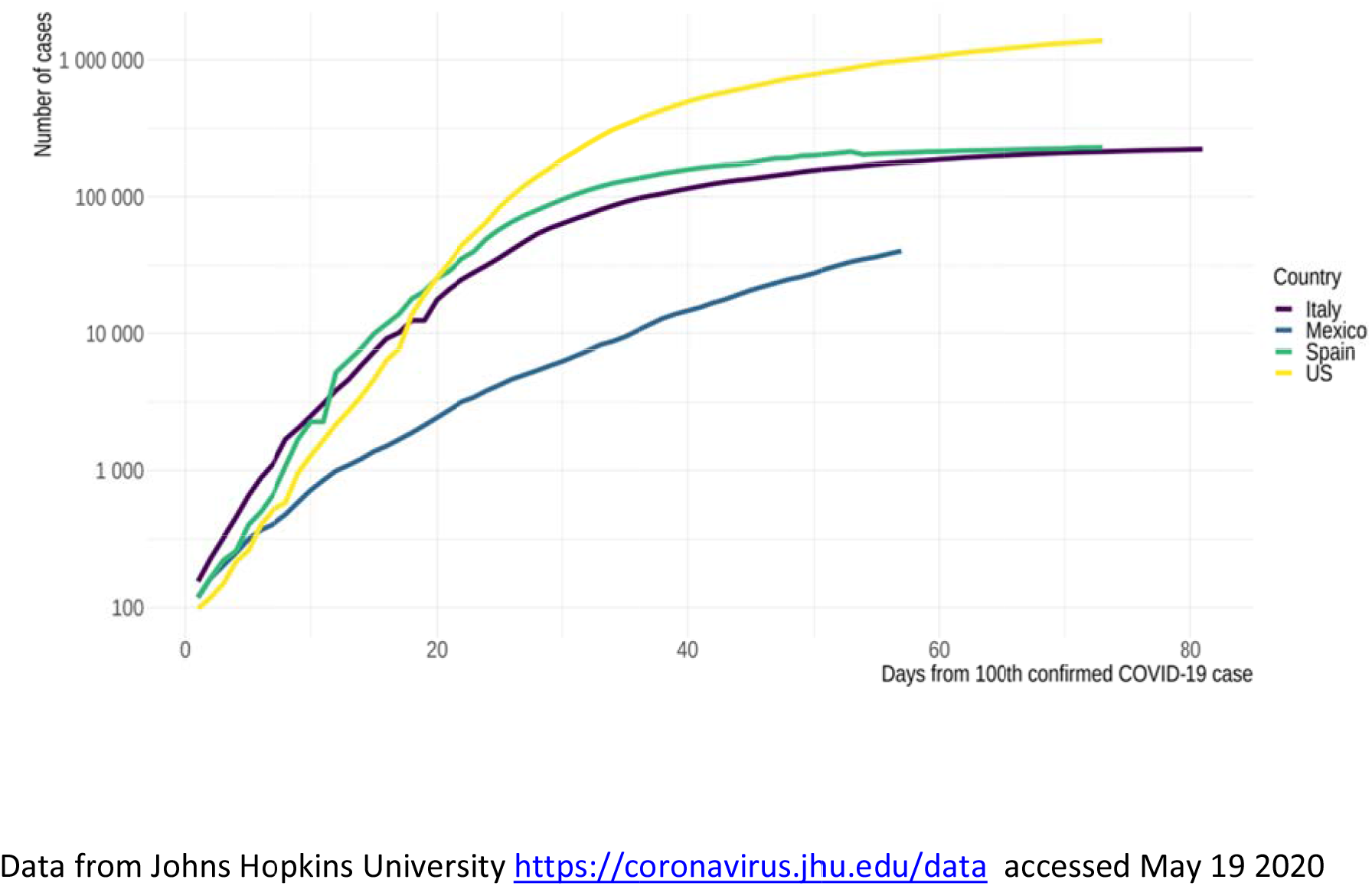
Comparison of transmission rate of Covid-19 in Mexico with that of other countries.

More significantly, up until early May there was no flood of patients overwhelming hospitals in Mexico. By May 14 it was reported (New York Times, 14 May) that in Mexico City, the city most severely affected, hospitals were full, but in other cities this had not occurred. This despite the fact that the Mexican population as a whole forms *a priori* a highly vulnerable population group: according to official Mexican health statistics, 76.8% of men and 73% of women are overweight or obese (Secretaria de Salud/INEGI 2018). Mexico moreover ranked 6th in the world for prevalence of diabetes in 2015 (Soto-Estrada et al. 2018). These represent two of the most important host factors contributing to COVID-19 morbidity and mortality (Muniyappa and Gubbi, 2020; Lighter et al. 2020). So although clearly there are data uncertainties, the evidence seems to indicate that though infections in Mexico are increasing, they are increasing at a slower rate than many other countries.

## Theory

It was discovered already in the late 19^th^ century that sunlight in the (ultra)violet-blue spectrum can kill micro-organisms (Hockberger 2000). UV-C radiation or UV_254_ (primary emission at 254 nm) is a potent killer of bacteria and viruses (Rauth 1965) and is used to sterilise water and in medical settings to sterilize equipment. UV-C is also dangerous to human beings, but in practice virtually all UV-C received naturally from the sun is filtered out by ozone in the stratosphere. The near-UV radiation that hits the ground is the portion of the spectrum in the range of 290 to 380 nm (UV-B and UV-A). Although UV-B and UV-A have a lower efficiency than UV-C, they act as natural viruscides in the environment (Rauth 1965) and their role in killing viruses on surfaces has been shown by experiments with artificial sunlight (Schuit et al. 2020). This is thus one route by which UV could potentially affect the rate of transmission of COVID-19.

However, there is another route by which UV may play a part in this. Humans have historically depended on the sun (in particular UV B photons) for vitamin D requirements (Holick, 2008), although modern (largely indoor) lifestyles have forced us to avoid Vitamin D deficiency by an array of other (mainly nutritional) means. Vitamin D is also a key element in the maintenance of healthy immune systems (Rondanelli et al 2018; Yamschchikov et al. 2009; Gunville et al. 2013). Hence it is plausible to suggest that UV may have a physiological effect on individual human beings which increases their level of immunity against COVID-19 infection and/or affects their susceptibility to (severe) disease, via production of vitamin D.

UV levels are much higher in the tropics than at higher latitudes, owing to the angle of the sun, and are higher in the southern hemisphere than in the northern at any equivalent latitude, owing to differences in the ozone layer. However, UV is also gradually filtered out as it passes through the atmosphere, so that other things being equal, higher levels of UV are received at higher altitudes, increasing at about 10-12% per 1000 meters of elevation (WHO 2018). Importantly, the amount received is strongly moderated by the scattering of UV radiation e.g. by cloud cover and atmospheric pollution. UV is measured locally on a scale of 1 to 15 by sensors, typically housed at airports or other major meteorological stations.

From an exploratory inspection of the data around mid-April it appeared that cities at higher elevations in Mexico had lower numbers of COVID-19 cases per 100,000 than cities at lower altitudes. We thus set out to test which geographical and meteorological variables might be correlated with this, focusing in particular on ultra-violet radiation (UV). In this paper we report on the extent to which: (a) the number of COVID-19 cases have been growing in different cities Mexico (i.e. city transmission rates) and (b) the rate at which deaths in these cities are accumulating, are correlated with the amount of UV these cities received before and during the period of transmission. We then discuss to what extent our data indicate that the mediating factor may be vitamin D.

## Method

We selected a sample of 45 cities (municipalities) over the whole of Mexico at low, medium and high altitudes, including larger and smaller cities, both coastal and inland, and cities with high and low levels of interaction with other parts of the country and the outside world (Figure 2). We chose the city, rather than state, level for the analysis because cities tend to have relatively small internal variations in temperature, humidity and UV while over a whole state these may vary considerably. The cities were selected at an early phase in the research before data on incidence of COVID-19 at municipal level was available; in other words we selected by geographical character, not by incidence of COVID, and we did not adjust the sample in any way at a later date. All cities had data on UV available from monitoring stations at their local airports^1^; this data was extracted from www.weatheronline.co.uk for the period 1 January to May 3 2020. We gathered data on daily maximum temperature (°C) and daily maximum relative humidity (i.e. % of water vapour in the air compared to the maximum the air at that temperature could hold) for the period from two weeks before the start of the transmission in each city up to May 3, from the UNAM site Interacción Océano Atmósfera/Fisicoquimica de la Atmósfera http://grupo-ioa.atmosfera.unam.mx/pronosticos/index.php.

**Figure 2:**
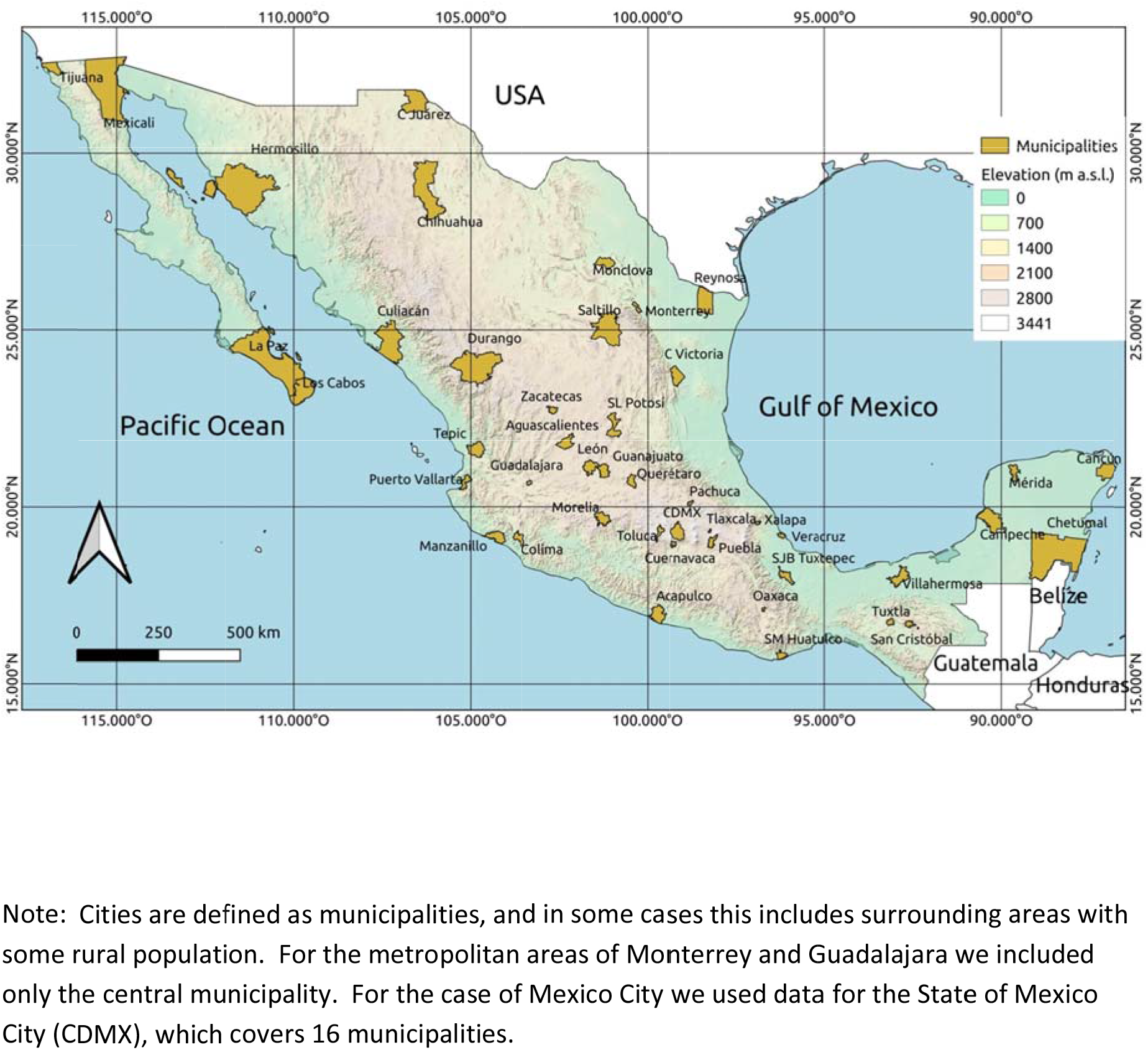
Map showing locations of cities included in the study and their elevations.

We extracted the weekly increment in confirmed COVID-19 cases and the number of COVID-19 ascribed deaths (both in cumulative terms, Tables 1 and 2) in each city, starting with the date of confirmation of the first case in the country (February 26 2020) up to May 3 2020, using official data from the Federal Secretariat for Health (Secretaria de Salud) (https://datos.COVID-19.conacyt.mx/). Although as noted this is probably a vast underestimate of actual cases, there is no reason to believe that this underestimate is significantly greater in some locations than in others^2^, so the relative level of infections in different cities should be adequately represented. The main explanatory variables (temperature, UV) increase as summer approaches, and COVID-19 cases are expected to increase exponentially whether or not they are related to the weather, so we analysed rates of transmission and rates of accumulated deaths over time (i.e. speed of increase in numbers of these in each city) rather than the absolute numbers of cases and deaths.

**Table 1:** Cumulative cases of COVID-19 in 45 cities across Mexico from 26 February to May 3 2020.

| City | Population<br>(million) | 1.3 | 8.3 | 15.3 | 22.3 | 29.3 | 5.4 | 12.4 | 19.4 | 26.4 | 3.5 | Cases/100,000<br>as of 3 May |
| --- | --- | --- | --- | --- | --- | --- | --- | --- | --- | --- | --- | --- |
| Aguascalientes | 0.9 |  |  | 2 | 9 | 41 | 47 | 57 | 126 | 207 | 296 | 32.9 |
| Tijuana | 1.9 |  |  |  | 11 | 59 | 225 | 412 | 682 | 966 | 1184 | 62.3 |
| Mexicali | 1 |  |  | 1 | 12 | 41 | 111 | 242 | 447 | 650 | 847 | 84.7 |
| Los Cabos | 0.08 |  |  |  | 3 | 14 | 58 | 135 | 169 | 191 | 208 | 260.0 |
| La Paz | 0.2 |  |  |  |  | 6 | 24 | 62 | 88 | 113 | 134 | 67.0 |
| Campeche | 0.2 |  |  |  | 1 | 4 | 5 | 8 | 12 | 22 | 43 | 21.5 |
| San Cristobal d/I Cs | 0.19 |  |  |  |  | 3 | 4 | 7 | 8 | 14 | 21 | 11.1 |
| Tuxtla | 0.5 |  |  | 1 | 2 | 7 | 15 | 18 | 32 | 51 | 77 | 15.4 |
| CDMX | 12.3 | 5 | 9 | 96 | 271 | 522 | 945 | 1786 | 3367 | 5728 | 8101 | 65.9 |
| Chihuahua | 0.88 |  |  | 1 | 3 | 4 | 13 | 25 | 48 | 98 | 171 | 19.4 |
| Ciudad Juarez | 1.3 |  |  | 1 | 6 | 18 | 50 | 93 | 162 | 276 | 402 | 30.9 |
| Saltillo | 0.8 |  |  | 1 | 6 | 10 | 18 | 26 | 29 | 37 | 51 | 6.4 |
| Monclova | 0.2 |  |  |  | 2 | 14 | 73 | 121 | 152 | 181 | 215 | 107.5 |
| Colima | 0.15 |  |  |  | 1 | 1 | 2 | 2 | 4 | 4 | 4 | 2.7 |
| Manzanillo | 0.11 |  |  |  |  | 1 | 2 | 5 | 9 | 15 | 19 | 17.3 |
| Durango | 0.65 |  |  | 3 | 4 | 5 | 5 | 6 | 6 | 20 | 35 | 5.4 |
| Guanajuato | 0.18 |  |  |  | 1 | 2 | 5 | 8 | 9 | 11 | 12 | 6.7 |
| Leon | 1.4 |  |  | 2 | 17 | 28 | 34 | 38 | 58 | 83 | 116 | 8.3 |
| Acapulco | 0.68 |  |  | 1 | 7 | 16 | 26 | 50 | 77 | 127 | 227 | 33.4 |
| Pachuca | 0.27 | 1 | 1 | 2 | 7 | 14 | 21 | 26 | 41 | 66 | 96 | 35.6 |
| Guadalajara | 1.5 |  |  | 6 | 27 | 38 | 52 | 62 | 75 | 102 | 137 | 9.1 |
| Puerto Vallarta | 0.22 |  |  | 1 | 2 | 6 | 11 | 19 | 32 | 53 | 89 | 40.5 |
| Toluca | 0.49 |  |  |  | 3 | 14 | 22 | 35 | 57 | 110 | 163 | 33.3 |
| Morelia | 0.6 |  |  |  | 9 | 16 | 22 | 24 | 31 | 42 | 51 | 8.5 |
| Cuernavaca | 0.33 |  |  |  | 2 | 8 | 19 | 26 | 54 | 145 | 251 | 76.1 |
| Tepic | 0.33 |  |  |  | 2 | 5 | 9 | 17 | 28 | 43 | 102 | 30.9 |
| Monterrey | 1.1 |  |  | 5 | 19 | 26 | 34 | 54 | 83 | 101 | 130 | 11.8 |
| Oaxaca | 0.26 |  |  | 1 | 2 | 9 | 12 | 12 | 13 | 24 | 28 | 10.8 |
| SJB Tuxtepec | 0.09 |  |  |  |  |  |  |  | 7 | 12 | 22 | 24.4 |
| Huatulco Sta Maria | 0.05 |  |  |  |  |  | 1 | 3 | 5 | 5 | 6 | 12.0 |
| Puebla | 1.4 |  |  | 5 | 15 | 57 | 126 | 187 | 262 | 362 | 484 | 34.6 |
| Queretaro | 0.8 |  |  | 6 | 11 | 23 | 36 | 53 | 77 | 100 | 129 | 16.1 |
| Cancun | 0.54 |  |  | 4 | 25 | 53 | 128 | 243 | 392 | 578 | 738 | 136.7 |
| Chetumal | 0.15 |  |  |  |  | 1 | 3 | 10 | 12 | 19 | 24 | 16.0 |
| San L Potosi | 0.82 |  |  | 2 | 20 | 32 | 39 | 49 | 55 | 69 | 103 | 12.6 |
| Culiacan | 0.67 |  |  |  | 3 | 33 | 115 | 235 | 433 | 628 | 831 | 124.0 |
| Hermosillo | 0.59 |  |  | 1 | 2 | 9 | 16 | 27 | 33 | 60 | 106 | 18.0 |
| Villahermosa | 0.35 |  |  |  | 4 | 39 | 91 | 160 | 341 | 581 | 808 | 230.9 |
| C Victoria | 0.27 |  |  |  |  | 2 | 3 | 8 | 25 | 92 | 124 | 45.9 |
| Reynosa | 0.61 |  |  |  |  | 2 | 11 | 15 | 23 | 34 | 45 | 7.4 |
| Tlaxcala | 0.09 |  |  |  |  | 2 | 5 | 7 | 8 | 16 | 33 | 36.7 |
| Veracruz | 0.55 |  |  | 1 | 4 | 6 | 14 | 44 | 101 | 212 | 375 | 68.2 |
| Xalapa | 0.52 |  |  |  |  | 3 | 6 | 7 | 12 | 15 | 19 | 3.7 |
| Merida | 0.89 |  |  | 5 | 28 | 46 | 82 | 130 | 204 | 304 | 418 | 47.0 |
| Zacatecas | 0.14 |  |  |  |  |  | 2 | 2 | 6 | 17 | 28 | 20.0 |

**Table 2:**
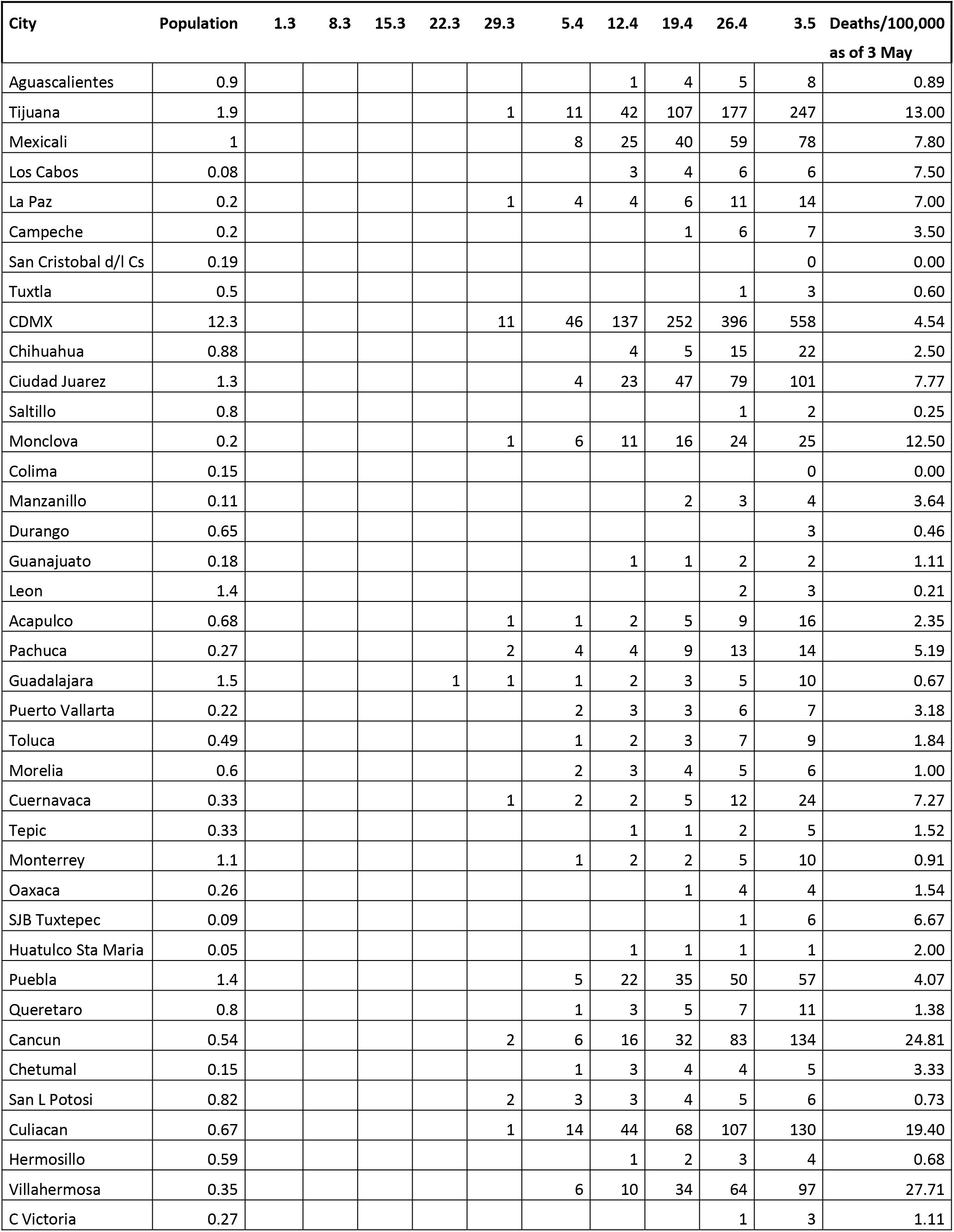
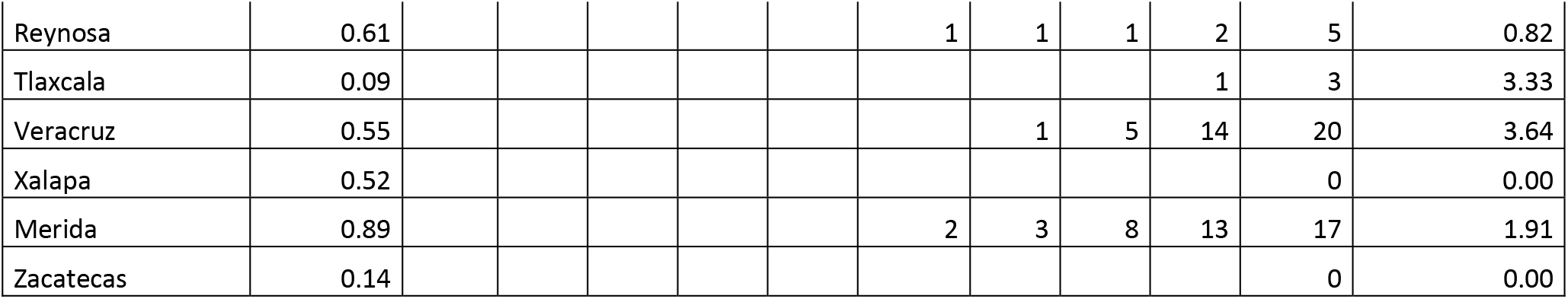
Cumulative COVID-19 ascribed deaths in 45 cities across Mexico, up to 3 May.

| City | Population | 1.3 | 8.3 | 15.3 | 22.3 | 29.3 | 5.4 | 12.4 | 19.4 | 26.4 | 3.5 | Deaths/100,000<br>as of 3 May |
| --- | --- | --- | --- | --- | --- | --- | --- | --- | --- | --- | --- | --- |
| Aguascalientes | 0.9 |  |  |  |  |  |  | 1 | 4 | 5 | 8 | 0.89 |
| Tijuana | 1.9 |  |  |  |  | 1 | 11 | 42 | 107 | 177 | 247 | 13.00 |
| Mexicali | 1 |  |  |  |  |  | 8 | 25 | 40 | 59 | 78 | 7.80 |
| Los Cabos | 0.08 |  |  |  |  |  |  | 3 | 4 | 6 | 6 | 7.50 |
| La Paz | 0.2 |  |  |  |  | 1 | 4 | 4 | 6 | 11 | 14 | 7.00 |
| Campeche | 0.2 |  |  |  |  |  |  |  | 1 | 6 | 7 | 3.50 |
| San Cristobal d/I Cs | 0.19 |  |  |  |  |  |  |  |  |  | 0 | 0.00 |
| Tuxtla | 0.5 |  |  |  |  |  |  |  |  | 1 | 3 | 0.60 |
| CDMX | 12.3 |  |  |  |  | 11 | 46 | 137 | 252 | 396 | 558 | 4.54 |
| Chihuahua | 0.88 |  |  |  |  |  |  | 4 | 5 | 15 | 22 | 2.50 |
| Ciudad Juarez | 1.3 |  |  |  |  |  | 4 | 23 | 47 | 79 | 101 | 7.77 |
| Saltillo | 0.8 |  |  |  |  |  |  |  |  | 1 | 2 | 0.25 |
| Monclova | 0.2 |  |  |  |  | 1 | 6 | 11 | 16 | 24 | 25 | 12.50 |
| Colima | 0.15 |  |  |  |  |  |  |  |  |  | 0 | 0.00 |
| Manzanillo | 0.11 |  |  |  |  |  |  |  | 2 | 3 | 4 | 3.64 |
| Durango | 0.65 |  |  |  |  |  |  |  |  |  | 3 | 0.46 |
| Guanajuato | 0.18 |  |  |  |  |  |  | 1 | 1 | 2 | 2 | 1.11 |
| Leon | 1.4 |  |  |  |  |  |  |  |  | 2 | 3 | 0.21 |
| Acapulco | 0.68 |  |  |  |  | 1 | 1 | 2 | 5 | 9 | 16 | 2.35 |
| Pachuca | 0.27 |  |  |  |  | 2 | 4 | 4 | 9 | 13 | 14 | 5.19 |
| Guadalajara | 1.5 |  |  |  | 1 | 1 | 1 | 2 | 3 | 5 | 10 | 0.67 |
| Puerto Vallarta | 0.22 |  |  |  |  |  | 2 | 3 | 3 | 6 | 7 | 3.18 |
| Toluca | 0.49 |  |  |  |  |  | 1 | 2 | 3 | 7 | 9 | 1.84 |
| Morelia | 0.6 |  |  |  |  |  | 2 | 3 | 4 | 5 | 6 | 1.00 |
| Cuernavaca | 0.33 |  |  |  |  | 1 | 2 | 2 | 5 | 12 | 24 | 7.27 |
| Tepic | 0.33 |  |  |  |  |  |  | 1 | 1 | 2 | 5 | 1.52 |
| Monterrey | 1.1 |  |  |  |  |  | 1 | 2 | 2 | 5 | 10 | 0.91 |
| Oaxaca | 0.26 |  |  |  |  |  |  |  | 1 | 4 | 4 | 1.54 |
| SJB Tuxtepec | 0.09 |  |  |  |  |  |  |  |  | 1 | 6 | 6.67 |
| Huatulco Sta Maria | 0.05 |  |  |  |  |  |  | 1 | 1 | 1 | 1 | 2.00 |
| Puebla | 1.4 |  |  |  |  |  | 5 | 22 | 35 | 50 | 57 | 4.07 |
| Queretaro | 0.8 |  |  |  |  |  | 1 | 3 | 5 | 7 | 11 | 1.38 |
| Cancun | 0.54 |  |  |  |  | 2 | 6 | 16 | 32 | 83 | 134 | 24.81 |
| Chetumal | 0.15 |  |  |  |  |  | 1 | 3 | 4 | 4 | 5 | 3.33 |
| San L Potosi | 0.82 |  |  |  |  | 2 | 3 | 3 | 4 | 5 | 6 | 0.73 |
| Culiacan | 0.67 |  |  |  |  | 1 | 14 | 44 | 68 | 107 | 130 | 19.40 |
| Hermosillo | 0.59 |  |  |  |  |  |  | 1 | 2 | 3 | 4 | 0.68 |
| Villahermosa | 0.35 |  |  |  |  |  | 6 | 10 | 34 | 64 | 97 | 27.71 |
| C Victoria | 0.27 |  |  |  |  |  |  |  |  | 1 | 3 | 1.11 |
| Reynosa | 0.61 |  |  |  |  |  | 1 | 1 | 1 | 2 | 5 | 0.82 |
| Tlaxcala | 0.09 |  |  |  |  |  |  |  |  | 1 | 3 | 3.33 |
| Veracruz | 0.55 |  |  |  |  |  |  | 1 | 5 | 14 | 20 | 3.64 |
| Xalapa | 0.52 |  |  |  |  |  |  |  |  |  | 0 | 0.00 |
| Merida | 0.89 |  |  |  |  |  | 2 | 3 | 8 | 13 | 17 | 1.91 |
| Zacatecas | 0.14 |  |  |  |  |  |  |  |  |  | 0 | 0.00 |

We log-transformed the data on accumulated COVID-19 cases and COVID-19 ascribed deaths in each city so that exponential growth rates were converted to linear ones. We calculated the slope of each of these growth rates to express per-city rates of transmission and death as single statistics (higher slope coefficients indicate faster rates of transmission and deaths for a given city). Data on temperature, relative humidity and UV radiation were then aggregated from their original daily resolution into different time periods. For each of these explanatory variables we calculated mean values for January and mean values since the first registered case of COVID-19 in each city. We then compared COVID-19 slope coefficients against the values of the explanatory variables using Pearson's r.

Data on vitamin D levels were extracted from a national nutrition and health survey carried out in 2018 (Secretaria de Salud/INEGI, 2018), which tested a small proportion of adult respondents for this and which included data on participants' municipalities of residence (including altitude).

The analysis was designed to examine the following hypotheses:

1. the speed of transmission of COVID-19 *is lower* at higher elevations
2. the speed of transmission of COVID-19 is related to temperature and relative humidity
3. the speed of transmission of COVID-19 *is lower* in cities with higher levels of UV radiation
4. The levels of UV in January *have a greater impact* on reducing rates of transmission than levels of UV during the period of transmission
5. the rate of increase in COVID-19 deaths *is lower* in cities with higher levels of UV radiation
6. the levels of UV in January *have a greater impact* on reducing rates of death than the level of UV during the period of transmission.

As regards the fourth and sixth hypotheses: as been noted above, two different kinds of relationship are possible: a sterilizing effect of UV in the environment, and a physiological effect of UV on individuals. Differences in the patterns of correlation could throw light on the mechanism involved, if any such relationship is shown. If the route is primarily by sterilization, then a clear negative relationship between rate of transmission and average UV during the period of transmission should be seen. If, on the other hand, the relationship is via a physiological effect, such as vitamin D, which builds up over time in the body, then it could be expected that differences in rates of infection would relate to differences in UV in the winter months previous to the infection. We selected the average UV in January to model this, since it is in this month that UV levels are lowest, especially in lower lying northern parts of the country. We stress that we are not out to prove that UV is the cause of either a sterilizing or a physiological effect, our aim is only to establish whether there is a correlation, and of what type. However any differences we find would be helpful in understanding whether vitamin D could be a mediating factor in the transmission of COVID-19.

## Results

The rates of transmission of COVID-19 in the cities sampled are shown in Figure 3. Since the epidemic started at different dates in different cities, data are shown from date of the first case (see also footnote 2).

**Figure 3.**
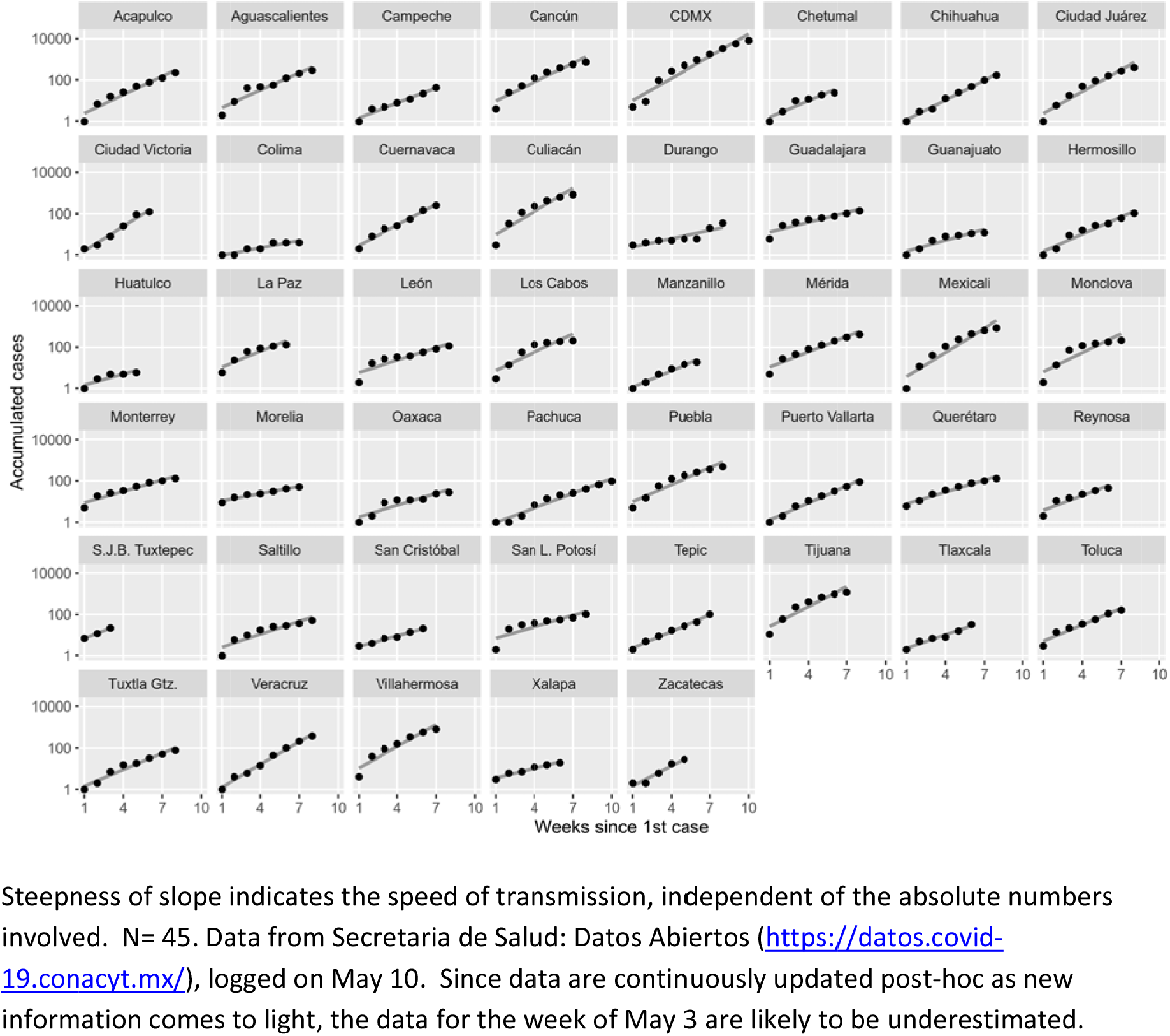
Transmission rates of Covid-19 up to May 3, in the 45 cities included in the study.

As regards hypothesis 1, we found that as expected, the rate of transmission is lower at higher elevations; this is a statistically significant result (Figure 4; Pearson's r = −0.365, p= 0.014), which indicates that about 13% (r^2^) of the variation in rate of transmission could be associated with differences in elevation. However, re hypothesis 2, neither temperature nor relative humidity during the period in which COVID-19 took hold and expanded (hypothesis 2) showed any relation to the rate of transmission (Supplementary materials Figures SM1 and SM2). Latitude was also ruled out as a factor, given that cities with highest transition rates (e.g. Tijuana, Culiacan, Mexico City, Villahermosa, Cancun) are found in the north, the centre and the south of the country, as are cities with the lowest rates (e.g. Monterrey, Saltillo, Guadalajara, Oaxaca, San Cristobal).

**Figure 4:**
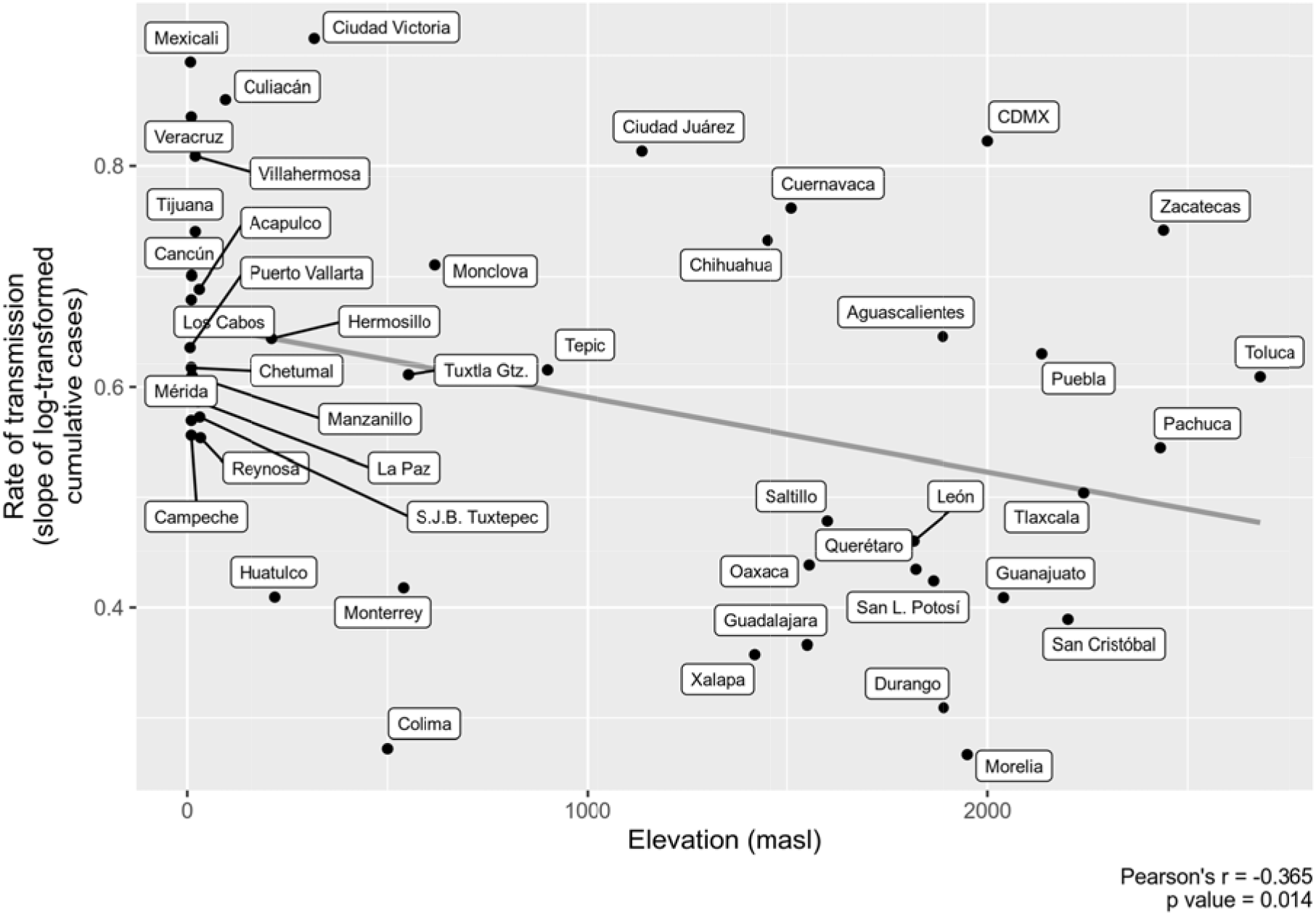
Relation of elevation to transmission rate of Covid-19.

For the case of hypothesis 3, we find a statistically significant lnegative trend between levels of UV during the transmission period and rate of transmission of COVID (Figure 5; r =.0.32, p = 0.032); cities with higher UV levels have slower rates of COVID transmission than cities with low UV levels.

**Figure 5:**
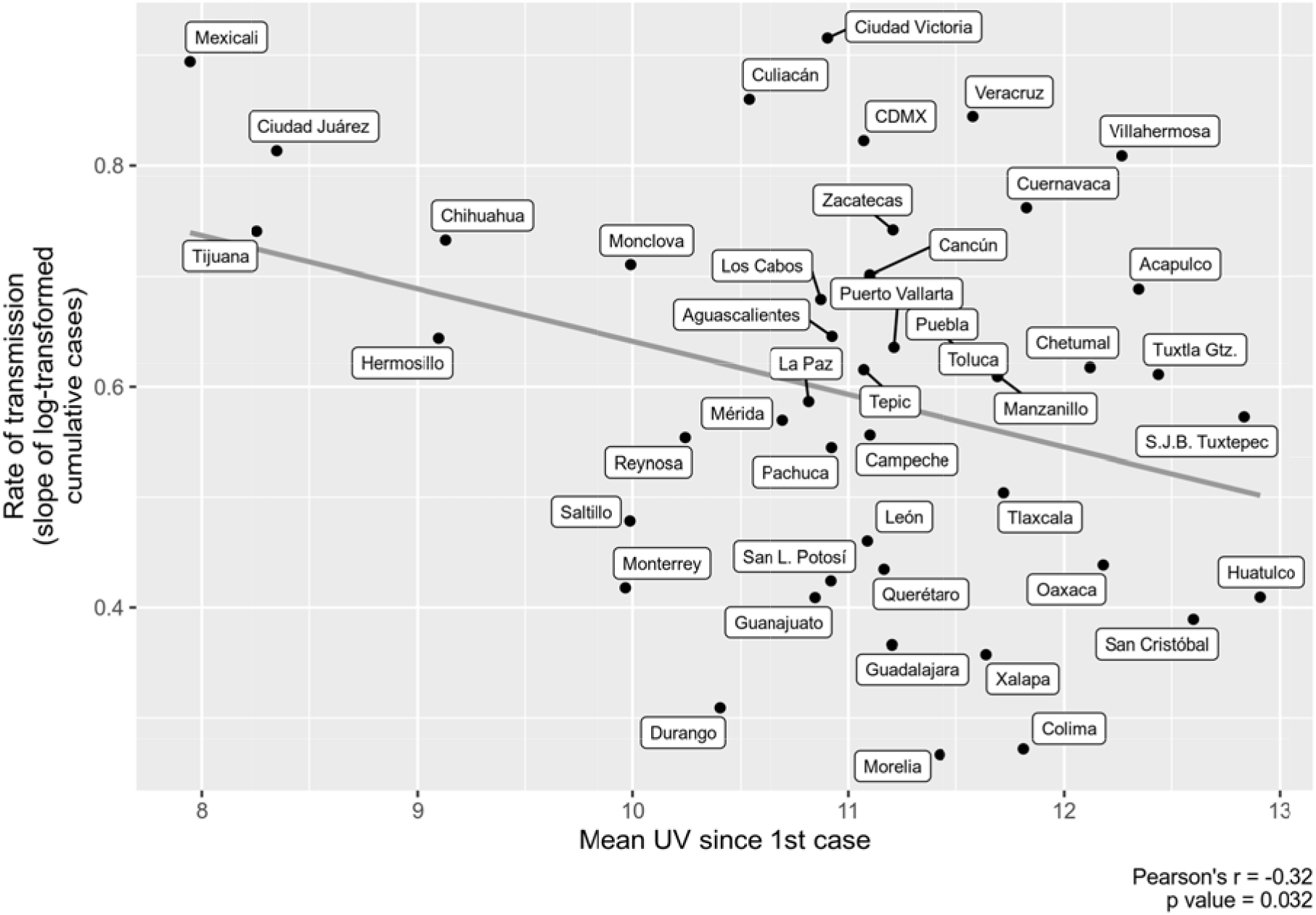
Relation of rates of transmission of Covid-19 to average UV during the period of transmission.

Interestingly, for hypothesis 4, our data show that there is a statistically significant negative relationship between average UV levels in January and speed of transmission of COVID from the start of the infection period (Figure 6; r= −0.369, p=0.014). This statistic is marginally more closely associated with the variation between cities in subsequent COVID-19 transmission than the level of UV during the period of transmission itself.

**Figure 6:**
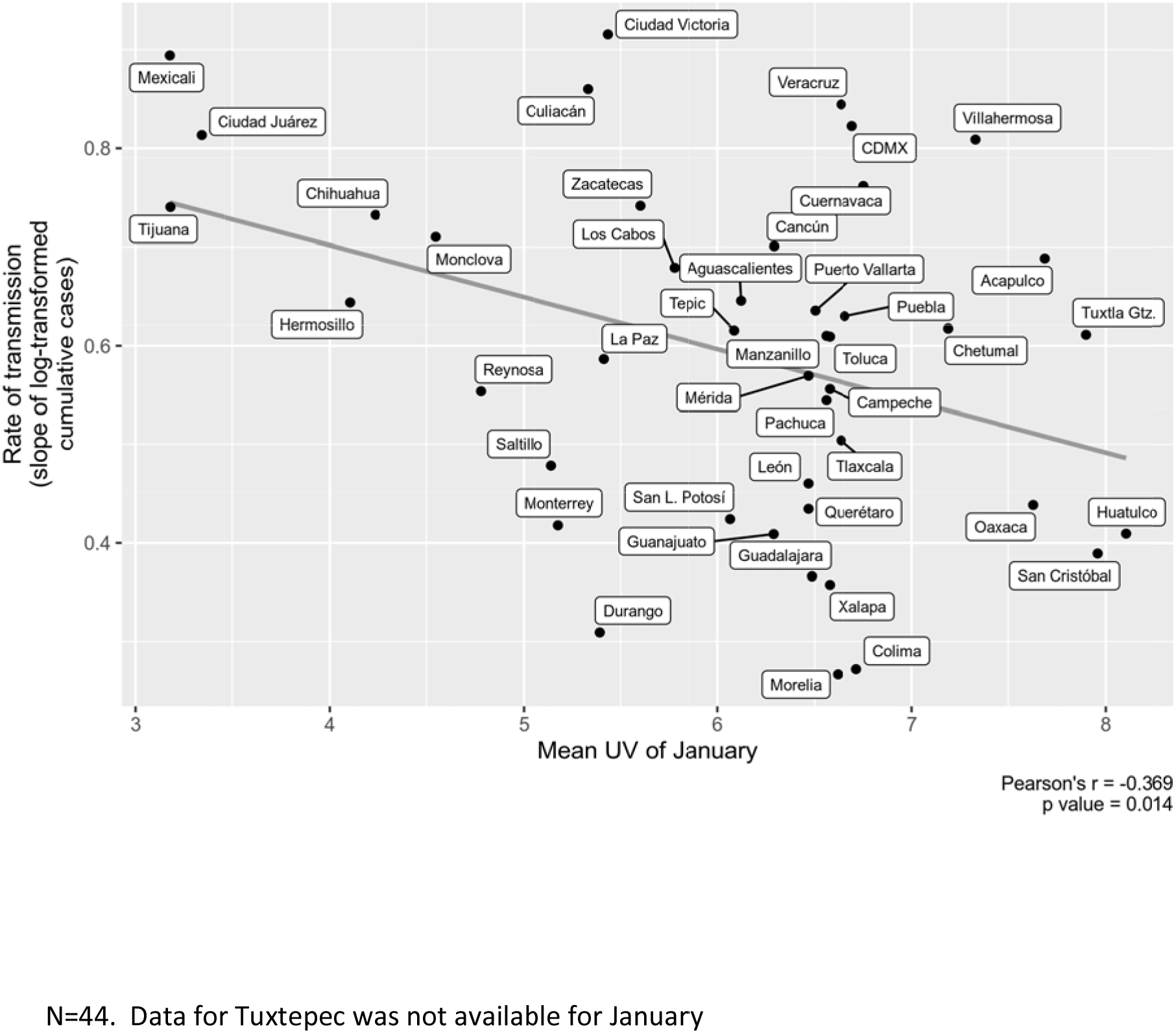
Relation of rate of transmission of COVID-19 to average UV in January.

Rate of cumulative deaths is presented in Figure 7 (each graph starts from the date of first death in the city), and as can be seen from the slopes, varies from city to city. Turning to hypotheses 5 and 6, there is a negative correlation between deaths and UV levels during the transmission period (Figure 8, r = −0.325, p = 0.07), which is marginally statistically significant, and also a negative correlation between deaths and January UV (Figure 9, r = −0.319, p = 0.081).

**Figure 7:**
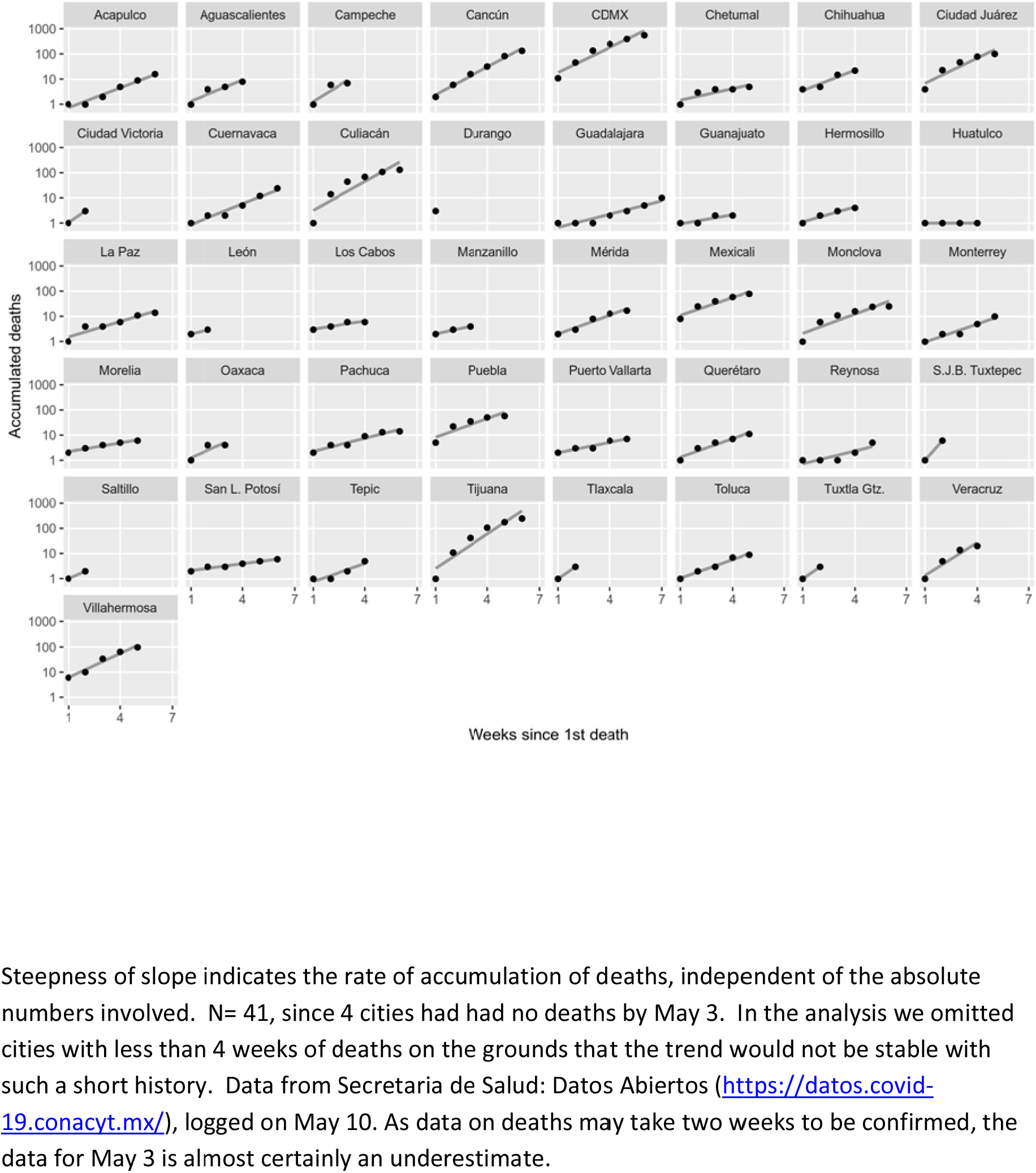
Rate of accumulation of COVID-19 ascribed deaths in the cities included in the study.

**Figure 8:**
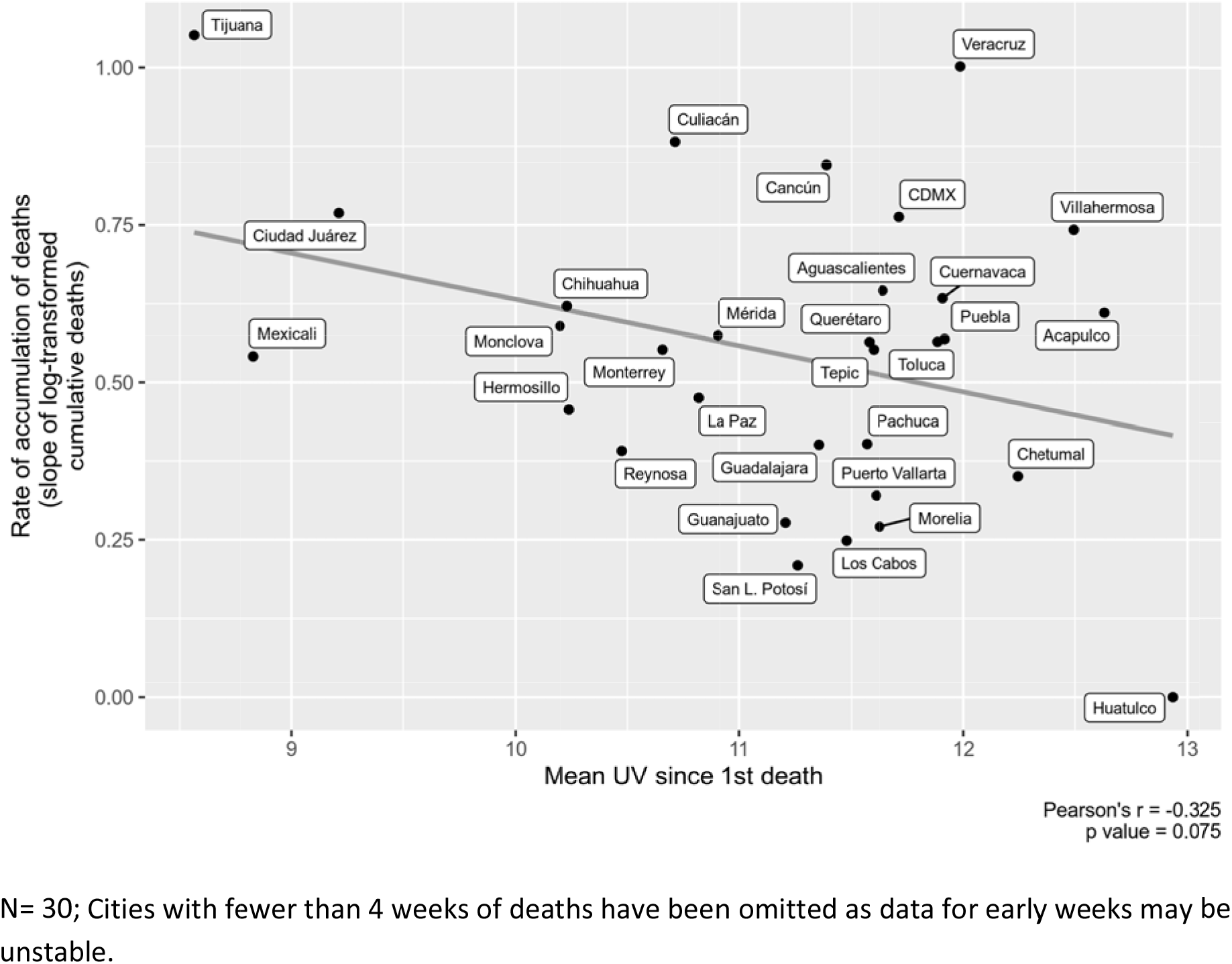
Relation of accumulation of COVID-19 ascribed deaths to average UV during the transmission period.

**Figure 9:**
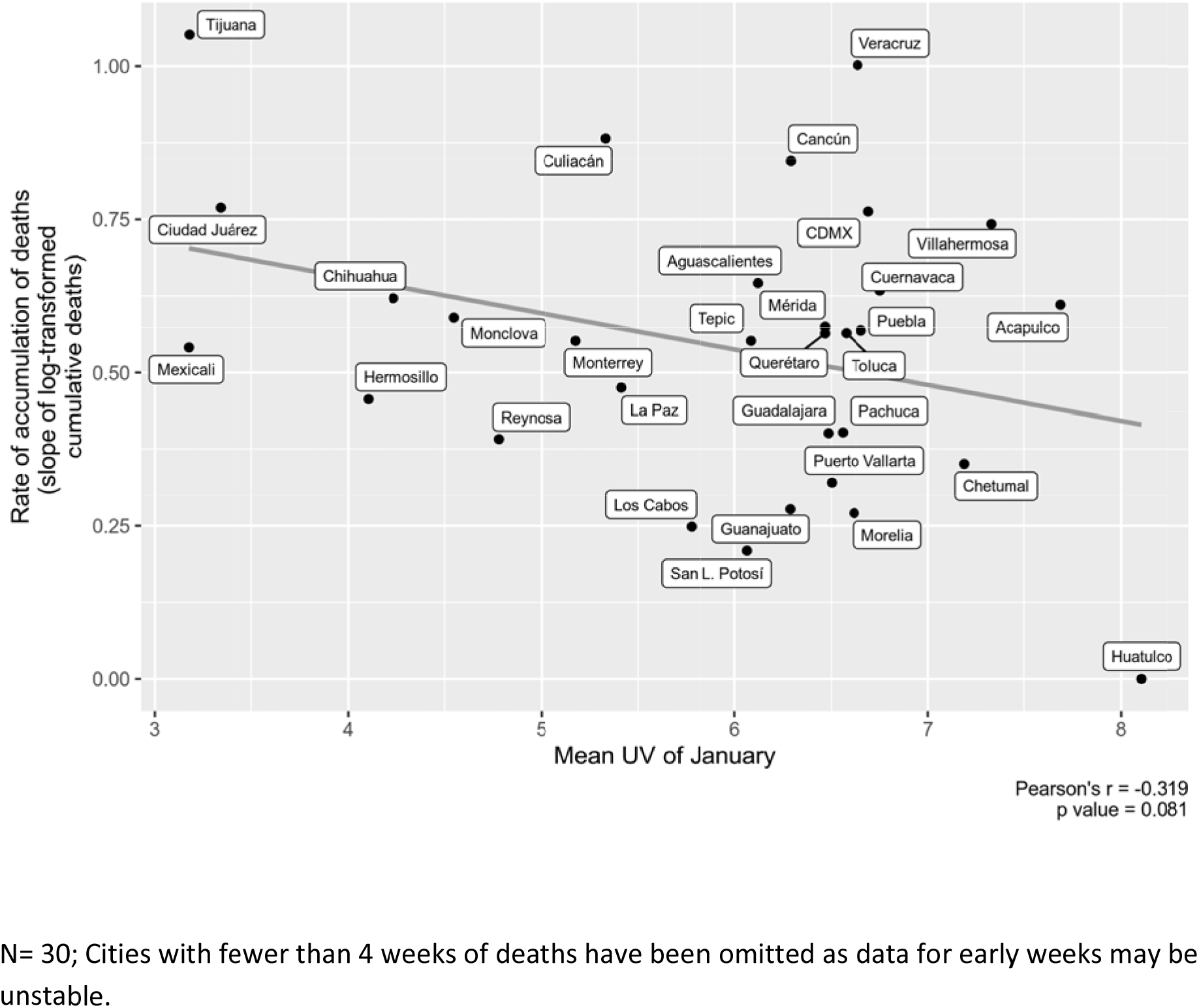
Relation of rate of accumulation of COVID-10 ascribed deaths to average UV in January.

Contrary to what we expected, altitude and UV levels in the Mexican cities are themselves not correlated (r = 0.09, p = 0.52), and indeed several coastal cities (though not all) have UV levels which are as high as those of many cities at 1500 meters and above. What the analysis shows is that UV and elevation *both* have an association with rates of transmission of COVID-19, but *independently* of each other. We therefore performed a multivariate regression, in order to determine how much of the variation can be explained when they are combined and which has a greater effect. This gave a highly significant (p= 0.0062) adjusted R^2^ of = 0.178, meaning that together they can account for about 18% of the variation in rates of transmission (Table 3). The standardized coefficients show that elevation has very slightly stronger effect than UV on transmission rates.

**Table 3:**
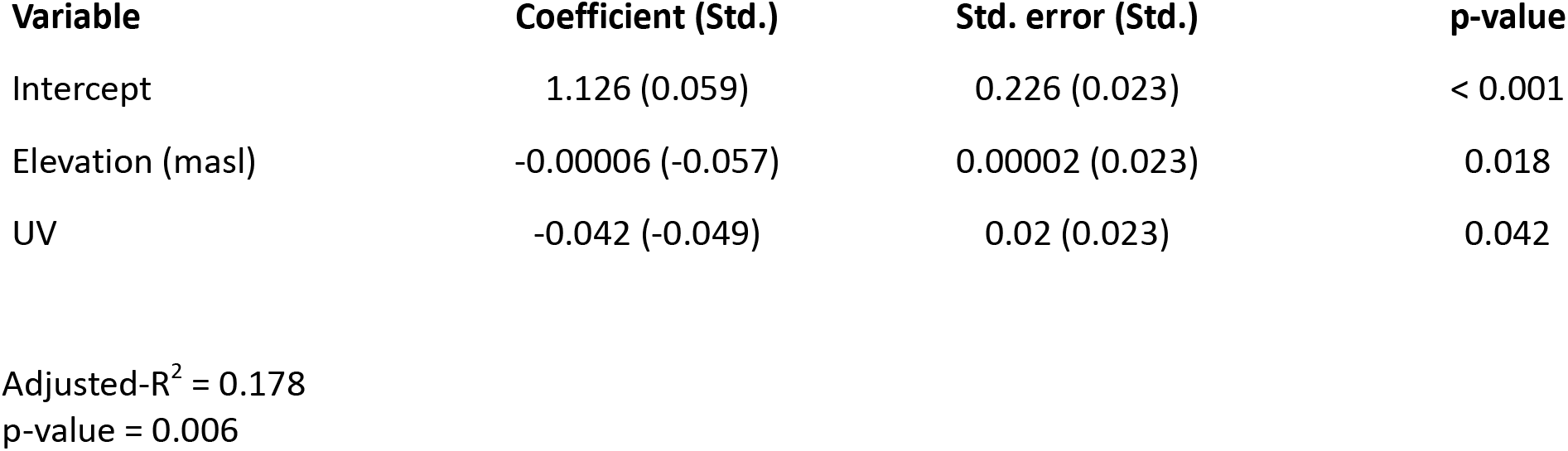
Coefficients for a multiple regression model between COVID-19 transmission rates vs. elevation and UV. Standardized coefficients and errors are shown in parentheses.

| **Variable** | **Coefficient (Std.)** | **Std. error (Std.)** | **p-value** |
| --- | --- | --- | --- |
| Intercept | 1.126 (0.059) | 0.226 (0.023) | < 0.001 |
| Elevation (masl) | -0.00006 (-0.057) | 0.00002 (0.023) | 0.018 |
| UV | -0.042 (-0.049) | 0.02 (0.023) | 0.042 |

Adjusted-R<sup>2</sup> = 0.178
p-value = 0.006

Finally, we analysed data from a national health survey carried out in 2018 (Secretaria de Salud/INEGI, 2018), which tested some (a small proportion) of the respondents for vitamin D levels, and we correlated this with elevation of the municipality of the respondent (Supplementary Materials Figure SM4). This showed that elevation is negatively correlated with vitamin D levels (r = −0.35, p = 00005). Unfortunately no data are available on the UV levels at these locations, since the 834 respondents who were tested for vitamin D were spread over 561 municipalities, many of them very small, and UV data are only available for larger cities with airports. For the same reason, insufficient vitamin D data were available for the individual cities included in our analysis to allow a meaningful assessment of whether mean vitamin D levels are associated with COVID-19 transmission levels.

## Discussion

Our results show that while temperature and relative humidity have no discernible association with the transmission rates of COVID-19 in Mexico from late February to early May, both altitude and UV levels are significantly associated with the transmission rates. This finding supports the idea that UV acts as a sterilizing force and reduces the concentration of viruses in the environment. Moreover we found a relationship between UV and rates of death over time, although with less statistical significance. We have not found a clear explanation for the evidently independent effect of elevation on transmission rates, although as we discuss below, the role of altitude may itself have a physiological explanation. Multivariate regression analysis showed that average UV during the transmission period and altitude together explain around 18% of the variation between cities as regards rates of transmission; many other factors are also clearly involved, as would be expected. Among these is the fact that UV only deactivates viruses by direct incidence, while a lot of transmission happens indoors or in places shaded from direct sunlight.

The findings on UV are largely in line with and complementary to those of other authors investigating differences in COVID-19 incidence between countries. Bäcker (2020) found that COVID-19 morbidity and mortality are both reduced in locations around the world with higher irradiance, citing solar angle (which is a close determinant of UV levels) as the most important factor in this. Kalippurayil et al. (2020) showed that across 64 countries included in their data set, one unit of increase in UV is associated with a 2.2% decline in cumulative COVID-19 deaths and a 1.9% decrease in daily rates of growth in numbers of cases. As stated, a major advantage of our own (national) dataset is that the confounding effects of factors such as human genetics, behavioural factors and public health and testing policy, will be markedly reduced compared to such international comparisons.

We also found that the association of UV with rates of transmission appears marginally stronger when looking at UV data in January 2020 (the month and a half before transmission started) than at UV data during the transmission period itself. This may reflect the pathway via which UV radiation affects transmission, i.e. through host physiology than direct sterilization of viral particles and thus potentially through the effects of UV on vitamin D levels.

There is considerable epidemiological evidence that vitamin D deficiency or insufficiency increases the risk of respiratory infectious diseases (Jolliffe et al. 2013; Jat 2017; Zisi et al. 2019). Evidence that vitamin D supplementation prevents respiratory infectious diseases is less clear-cut, presumably reflecting heterogeneity in (definitions of) baseline vitamin D deficiency and dosing regimens, although the most recent meta-analysis to date does suggest a beneficial effect (Martineau 2017). Mechanisms of protection by vitamin D have been suggested to include both induction of the antimicrobial peptides hCAP-18/LL-37 and human β-defensin 2 (Wang 2004), which also have antiviral properties (Kota 2008, Barlow 2011, Currie 2013), and the dampening of inflammatory immunopathology during infection (reviewed in Greiller & Martineau 2015 and Zdrenghea et al. 2017). The latter might be of particular pertinence in the context of COVID-19, where immunopathology is considered to play a large role in late-stage disease (van de Veerdonck et al. 2020).

Daneshkhah et al. (2020) have signalled the importance of vitamin D in reducing the death rate in COVID-19. Based on clinical data from COVID patients in China, they conclude that those with low vitamin D levels are more likely to suffer cytokine storm during progressive disease, which is thought to be the immediate cause of death in many severely ill COVID patients. The role of vitamin D in reducing severe manifestations of COVID-19 is also proposed in a very recently published study which suggests that differences in national mean vitamin D levels secondary to supplementation policies are a major factor behind the variation in COVID-19 mortality rates across countries in Europe (Laird et al. 2020). These authors use their findings to urge governments to revise current advice on vitamin D supplements in the light of their potential use in lowering the likelihood of severe reactions to SARS-CoV2 infection, also discussed by Davies et al. (2020), and it has been reported in the press (La Voz de Michoacan, 19 May) that severely ill patients are being treated with injected vitamin D in Spain. Although many studies which deal with the influence of vitamin D on COVID-19 focus on its effects in reducing severe immunopathology in late-stage disease, rather than its role in enhancing viral clearance early during infection, in practice the effect of both mechanisms would overlap, resulting in lower morbidity across the complete clinical spectrum of COVID-19. Moreover, these effects might even appear to lower transmission rates, since if vitamin D helps to suppress the more serious effects of COVID-19, it follows that more people who become infected develop only mild symptoms and may never present for testing and be registered as confirmed cases.

As regards the link between levels of UV exposure and vitamin D levels: when UV levels are low (< 4 on an index of 0-15), as in northern winters, it is known that many people have vitamin D deficiency (Cannell et al. 2006); typically bodily Vitamin D levels rise in the summer. The winter effect may be compounded by the fact that people do not go out very much and wear more clothing. However, there are differences at individual level in ability to manufacture vitamin D, as well as between different population groups; for example, older people in general have reduced ability to do this (Boucher 2012, van der Wielen 1995). Moreover melatonin greatly affects the body's ability to absorb UV. In this regard, it is noted that recent studies have indicated that in the UK, people from ethnic minorities are much more susceptible to COVID-19 than the Caucasian population. After correcting for income, underlying health and other social conditions, black men are twice as likely to die of COVID-19 as Caucasian men of a similar age, and men of Pakistani or Bangladeshi origin, 1.8 times (UK Office of National Statistics, 7 May 2020; Aldridge et al. 2020b). There is as yet no established explanation for these differences. Physiological differences, such as relative ability to produce Vitamin D at given levels of UV radiation, thus remain a possible cause.

For the case of Mexico, although we had access to data on vitamin D levels from a subset of subjects screened in a nationwide public health survey, there were insufficient data available per city in our study to meaningfully assess correlations with either UV levels or COVID-19 transmission. We found a *negative* association between vitamin D levels and altitude. This is in line with studies on children living at different altitudes in the Andes (Hirschler 2013 & 2019, Teran 2017). The proposed explanation for this is that children at altitude wear more clothing and spend less time outdoors due to the colder and windier climate; poorer nutrition (particularly access to vitamin D-rich foods such as fish and meat) may also play a role. The same might hold true in the overall population of the Mexican national public health survey (which included inhabitants of poor rural communities), though not necessarily for the population of the cities included in our COVID-19 data set. But as our data clearly show that, contrary to expectation, UV is not correlated with elevation, it would be surprising had we found a strong positive relationship of vitamin D with altitude. Without knowing the actual average vitamin D levels in the populations of the cities we studied, we can therefore only speculate on whether the observed association between UV levels and COVID-19 transmission is mediated by vitamin D.

As regards the interesting association that altitude appears to have with transmission rates, quite independently of UV, it has recently been proposed that COVID-19 transmission may be reduced physiologically as a result of the lower oxygen levels at high elevations, through hypoxia-mediated down-regulation of the angiotensin-converting enzyme 2 (ACE2), which forms the primary binding site of the causative virus SARS-CoV2 (Arias-Reyes et al. 2020). In other words, down-regulation of ACE2 expression on cells in the lung could slow down multiplication of the virus. Clearly further research is needed to follow up this interesting lead.

We were not able to test various other hypotheses which could also explain variations in COVID-19 transmission rates. For example, it is possible that high levels of air pollution predispose populations to the disease through lung damage; particulate matter is often cited in this context (e.g. Zanobetti et a. 2000) although ozone has also been suggested (Alipio 2020). Particulate matter (PM10 and PM2.5) may also promote spread of the virus in the air or keep it in suspension for longer. Although there is extensive data for air pollution in Mexico City, there is very little for other cities (see Supplementary Materials Figure SM4), and we were not able to test for this. Smoke from indoor cooking fires may also play a part, since a large proportion of the rural population of Mexico (although admittedly not the urban population, represented by our sample) still relies on woodfuel as the primary energy source (Masera et al. 2020).

Finally we note that our data covered only the first ten weeks of the outbreak of COVID-19 in Mexico and that clearer patterns may emerge in due time.

## Conclusions

Our study indicates that in addition to the sterilizing effect, there is a link between UV radiation and the rates of transmission and mortality of COVID-19, potentially via the production of vitamin D. It adds strength to calls by other researchers for biomedical studies to urgently investigate the link between vitamin D and susceptibility to COVID-19, both mechanistically and in well-designed randomised clinical trials. Our results also suggest that the independent association of elevation to COVID-9 needs further investigation.

## Data Availability

All data were obtained from sources that are publicly accessible (URLs are cited in the paper). We are willing to share the database we created from these data, on request.

## Acknowledgements

The authors would like to thank John Skutsch, Robin Reifel, Emily Sturdivant, Cyndie Katz and Elizabeth Waterston for comments and suggestions.

## Footnotes

1 A possible confounding factor is that airports are usually located at some distance from the city, so that their UV readings may not be accurate for the city centre itself. However, there is no other source for UV data so we have to accept this potential source of error.

2 A possible confounding factor here is that cities whose transmission starts were one or two weeks later than the others might have had time to establish more systematic testing and reporting procedures which might bias the reliability of data. The vast majority of our sample however reported first cases in the weeks ending 8 or 15 March (Table 1). We were not able to establish how rapidly each municipality introduced testing but Mexico has a public health system controlled at Federal level and we think it unlikely that there have been major differences in speed of response across different cities.

